# Perceived versus proven SARS-CoV-2 specific immune responses in health care professionals

**DOI:** 10.1101/2020.05.12.20094524

**Authors:** Georg M.N. Behrens, Anne Cossmann, Metodi V. Stankov, Torsten Witte, Diana Ernst, Christine Happle, Alexandra Jablonka

**Affiliations:** Department for Rheumatology and Clinical Immunology, Hannover Medical School, Hannover, Germany; German Center for Infection Research (DZIF), partner site Hannover-Braunschweig, Germany; Department of Pediatric Pneumology, Allergology, and Neonatology, Hannover Medical School, Hannover, Germany; German Center for Lung Research, Biomedical Research in End Stage and Obstructive Lung Disease/BREATH Hannover

**Keywords:** COVID-19, SARS-CoV-2, Immunoglobulin, IgG, IgA, health care worker, ELISA, Seroprevalence, Diagnostics, health care professionals

## Abstract

There have been concerns about high rates of thus far undiagnosed SARS-CoV-2 infections in the health care system. The COVID-19 Contact (CoCo) Study follows 217 frontline healthcare professionals at a university hospital with weekly SARS-CoV-2 specific serology (IgA/IgG). Study participants estimated their personal likelihood of having had a SARS-CoV-2 infection with a mean of 20.9% (range 0 to 90%). In contrast, anti-SARS-CoV-2-IgG prevalence was about 1-2% at baseline. Regular anti-SARS-CoV-2 IgG testing of health-care professionals may aid in directing resources for protective measures and care of COVID-19 patients in the long run.

## Brief Report

Despite growing access of broadly available testing systems, uncertain rates of asymptomatic infections have raised concerns about a potentially high rate of thus far undiagnosed SARS-CoV-2 infections, particularly in frontline medical staff (1). To prevent the breakdown of health-care systems during the current pandemic, the protection of medical personnel and patients from contracting a SARS-CoV-2 infection is central (2).

Consent finding for case definition, COVID-19 diagnosis in suspected cases, and scaling up of suitable diagnostic systems have been challenging since the start of the pandemic. Realtime polymerase chain reaction (PCR) based throat swab testing was rapidly developed and has helped in ascertainment and tracking of the SARS-CoV-2 outbreak (2). However, sensitivity of PCR based testing, which is thus far only applied routinely for symptomatic patients, crucially depends on timing and type of respiratory sampling and led to false negative rates of up to 70% during the early phase of the pandemic (3,4). Serological testing for SARS-CoV-2 specific immunoglobulins (Ig) is relatively easy, inexpensive, and critical for epidemiological studies. SARS-CoV-2 specific B cell responses appear to correlate to disease severity with rising antibody titers typically between 5 to 10 days and fully positive rates at about 18 days after symptom onset (5). As such, serological testing can be helpful in suspected cases with negative PCR results and in identification of asymptomatic infections (6).

We initiated the COVID-19 Contact (CoCo) study to weekly monitor SARS-CoV-2 specific serology (IgA/IgG) in frontline healthcare professionals (HCP) in combination with a questionnaire about respiratory symptoms and risk perception. As testing system, we employed a semiquantitative ELISA [EUROIMMUN Medizinische Labordiagnostik, Lübeck, Germany – CE certified version: specificity 99.0 %, sensitivity 93.8 % after day 20 according to manufacturer (5)]. We confirmed the specificity in a set of 156 sera from non-European refugees and migrants (7) collected in 2015 as negative controls (mean age 31.6 years, range 18 – 67 years, 78% male). All but one tested negative for SARS-CoV-2 IgG (specificity 99.3%) and two out of 90 tested equivocal positive for IgA (specificity 97.8%). Serum of 18 patients after recovery of PCR-confirmed COVID-19 served as positive controls or to generate a standard curve (mean age 44.8 years; mean duration of symptoms 11.8 days, range 3-35 days; mean time since start of symptoms 30.4 days, range 21-61 days). 16/18 tested positive (n=1 equivocal positive) for SARS-CoV-2 IgG (sensitivity 90%) and 15/18 positive for SARS-CoV-2 IgA (sensitivity 85.7%). Interestingly, the duration of symptoms as a surrogate for disease severity correlated significantly with the IgG ratio (extinction of sample to calibrator ratio) of the SARS-CoV-2 IgG ELISA (Figure A).

**Figure:**
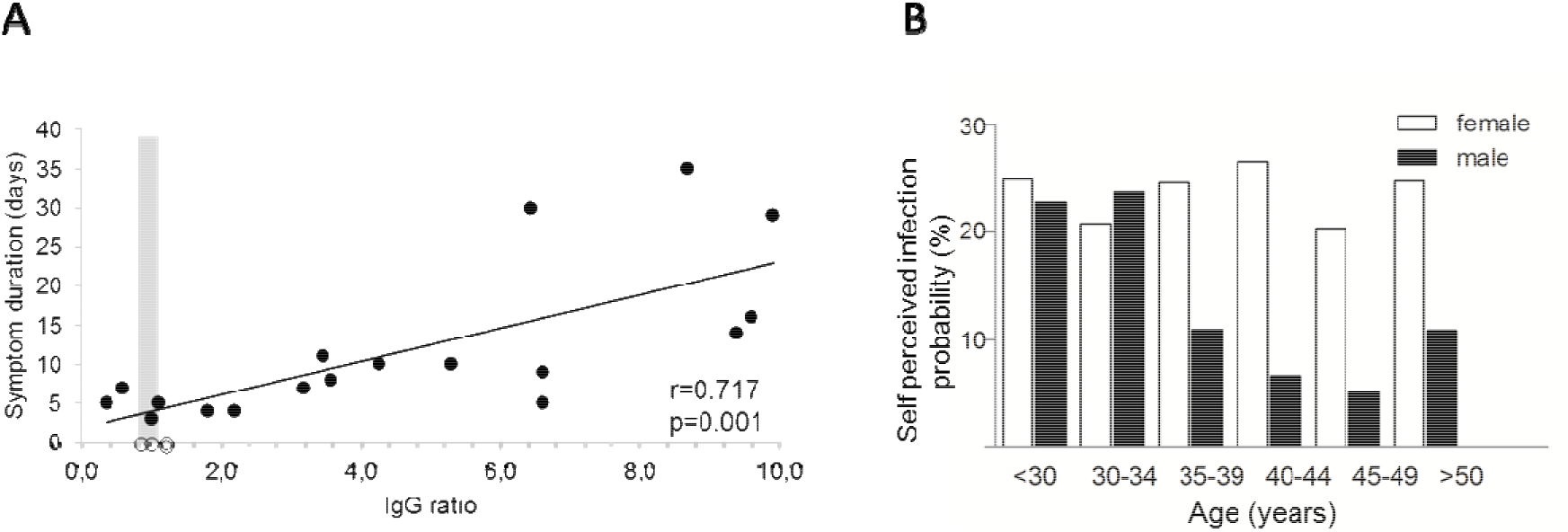
(A) Anti-SARS-CoV-2 IgG ELISA results. PCR-confirmed COVID-19 cases are depicted as black dots, health care professionals depicted as open dots (for which symptoms were not considered). The grey zone (0.8-1.1 ratio) represents the range with equivocal ELISA results. (B) Differences in self perceived probability for SARS-CoV-2 infection in relation to sex and age.

Between March 23rd and April 17th, n=217 HCP from emergency rooms, infectious and pulmonary disease wards, ICUs, pediatric departments and other units involved in COVID-19 patient care at our university hospital were included in the study. The mean age of participants was 36.5 years (range 18 to 63 years), and 65% were female. Most of them worked as physicians (53.5%), nurses (27.6%), or medical assistants (9.2%). The majority of participants had direct contact to patients with infectious respiratory diseases working in the emergency department (40.1%), general ward (31.8%), or outpatient departments (13.8%). At baseline, 1.6% of included personnel reported to have visited regions with high SARS-CoV-2 prevalence as defined by the German national institute of public health (Robert Koch Institute (8)), 16.1% reported to have had contact to confirmed COVID-19 cases, and more than one third (39.2%) to have had contact to suspected COVID-19 cases. 45.2% of HCP reported to suffer from at least one respiratory symptom of any severity, and 29.0% reported to have had a respiratory infection during the past two weeks.

Upon enrollment, study participants were asked to estimate their personal likelihood of having had a SARS-CoV-2 infection. Only 12% of the n=201 study participants, who answered this question, rated a zero chance of having already had contracted SARS-CoV-2. Strikingly, the mean percentage of self-perceived positive SARS-CoV-2 infection status was as high as 20.9% (range 0 to 90%). Male participants rated their infection risk lower than female participants (mean 16.2% vs. 23.7%, F=4.4, p=0.02) and older subjects reported lower infection probabilities as compared to younger participants (Pearson -0.33, p=0.004) (Figure B). Reported contact to confirmed or suspected COVID cases did not have a significant impact on perceived probability of infection.

In contrast to the high percentage of self-perceived SARS-CoV-2 infection, only two of n=217 tested frontline HCP showed a clearly positive reaction in the ELISA, and two displayed equivocal positive results according to the manufacturer’s interpretation. Both positive results were about 20-fold lower as compared to one patient with severe COVID-19 (Figure A). The majority of participants (n=214) had no evidence of anti-SARS-CoV-2-IgG. Anti-SARS-CoV-2-IgA was positive/equivocal positive in n=9 and n=10 subjects respectively, and combined IgG and IgA positive/equivocal positive in three subjects. Altogether, anti-SARS-CoV-2-IgG prevalence was in the range of 1-2%.

Our data on SARS-CoV-2-IgG is only partially representative for our university hospital, not fully representative for other clinics, and we do not know the source of infection in anti-SARS-CoV-2 IgG positive HCP. However, the gap between perceived risk and evidence for an infection is most likely a phenomenon in many health care settings. Additionally, we have only limited information about the full validity of anti-SARS-CoV-2 serology tests for screening. In a setting with low COVID-19 prevalence, the use of the spike protein S1 to screen for anti-SARS-CoV-2 IgG may be suboptimal, and testing for antibodies against e.g. the receptor-binding domain of SARS-CoV-2 could increase sensitivity. Interestingly, given the significant association between disease severity and our anti-SARS-CoV-2-IgG in our ELISA, we hypothesize that asymptomatic seroconversions could lead to numerous equivocal positive ELISA results (Figure A), which still may represent neutralizing activity (9). Such data may be difficult to interpret in cross-sectional studies and our longitudinal study design combined with neutralization assays will be informative about the magnitude of ELISA result changes over time. Finally, a matter of debate remains whether serological tests can also inform about COVID-19 specific immunity. Preliminary studies in rhesus macaques suggest that reinfection does not occur after survival of COVID-19, supporting the notion of at least temporary immunity after primary infection (10).

Taking these limitations into account, our data points towards a currently low rate of SARS-CoV-2 specific IgG in HCP in Northern Germany hospitals, where no overflow of COVID-19 patients has challenged the healthcare system so far and confirmed outbreaks are limited. This is in stark contrast to the relatively high rate of self-estimated SARS-CoV-2 infections of our hospital’s frontline HCP and strikingly different from currently high infection rates in medical personnel from Italian regions (1). Also, our data show that personal risk perception correlates to age and sex, which should be taken into account when advising hospital staff on protective measures against COVID-19. Regular anti-SARS-CoV-2 IgG testing of health-care workers may aid in monitoring the pandemic, assessing the quality of immune responses, and directing resources to assure COVID-19 care in the long run.

## Data Availability

Individual deidentified participant data that underlie the results will be shared. The study protocol will be available 9 -36 months after publication with researchers who propose a methodically sound proposal, proposals should be directed to

## Ethics approval and consent to participate

The here presented analyses were approved by local authorities (Data Security Management and Institutional Review Board of Hannover Medical School), approval number 8973_BO_K_2020). Informed consent was obtained from all participants.

## Consent for publication

Not applicable

## Availability of data and materials

The datasets used and/or analysed during the current study are available from the corresponding author on reasonable request.

## Competing interests

AJ reports grants and personal fees from Novartis, grants and personal fees from Abbvie, grants and personal fees from Gilead, personal fees from Roche, outside the submitted work; TW reports grants and personal fees from Novartis, grants and personal fees from Abbvie, personal fees from Gilead, personal fees from Chugai, personal fees from Sanofi-Aventis, non-financial support from Aesku.Diagnostics, outside the submitted work; DE Dr. Enst reports grants and personal fees from Novartis, grants and personal fees from Abbvie, grants and personal fees from Gilead, personal fees from Sanofi Aventis, personal fees from GSK, outside the submitted work; Other authors have nothing to disclose.

## Funding

There has been no external funding for this project.

## Authors’ contributions

GB, CH, AJ designed and managed the project, analyzed the data and drafted the manuscript. AC managed proband inclusion and designed the workflow. MS performed the experiments and designed the laboratory workflow, DE and TW were involved in planning and supervised the project. All authors discussed the results and approved the final manuscript.

## Acknowledgements

We thank the study site coordinators and participants of the CoCo Study group.

**Abbreviations**
COVID-19: coronavirus disease 19
ELISA: Enzyme-linked Immunosorbent Assay
IgG: immunoglobulin G
PCR: real-time polymerase chain reaction
SARS-CoV-2: Severe acute respiratory syndrome coronavirus 2

## Notes

### Clinical Trial

DRKS00021152

### Funding Statement

No external Funding for this work was received.

